# Cost-effectiveness of intensive care for hospitalized Covid-19 patients: experience from South Africa

**DOI:** 10.1101/2020.10.30.20222802

**Authors:** SM Cleary, T Wilkinson, CR Tamandjou Tchuem, S Docrat, GC Solanki

**Author notes:** Corresponding Author: Susan Cleary Correspondence Address: Health Economics Unit, School of Public Health and Family Medicine, University of Cape Town.

## Abstract

**Background:** Amidst the shortages of critical care resources in the public sector resulting from the COVID-19 pandemic, the South African Government embarked on an initiative to purchase critical bed capacity from the private sector. Within an already under-funded public health sector, it is imperative that the costs and effects of potential interventions to care are assessed and weighed against the opportunity costs of their required investment.

**Objective:** To assess the cost-effectiveness of ICU management for admitted COVID-19 patients across the public and private health sector in South Africa.

**Methods:** Using a Markov modelling framework and a health system perspective, the costs and health outcomes of inpatient management of severe and critical COVID-19 patients in (1) general ward and intensive care (GW+ICU) and (2) general ward only were assessed. Disability adjusted life years (DALYs) were evaluated as health outcomes, and the cost per admission from public and private sectors was determined. The models made use of four variables: mortality rates, utilisation of inpatient days for each management approach, disability weights associated to the severity of the disease, and the unit cost per general ward day and per ICU day in public and private hospitals. The unit costs were multiplied by utilisation estimates to determine the cost per admission. DALYs were calculated as the sum of years of life lost (YLL) and years lived with disability (YLD). An incremental cost-effectiveness ratio (ICER) representing the difference in costs and health outcomes of the two management strategies - was calculated and compared to a cost-effectiveness threshold to determine the value for money of ICU management.

**Results:** A cost per admission of ZAR 75,127 was estimated for inpatient management of severe and critical COVID-19 patients in general wards only as opposed to ZAR 103,030 in GW+ICU. DALYs were 1.48 and 1.10 in the general ward only and GW+ICU, respectively. The ratio of difference in costs and health outcomes between the two management strategies produced an ICER equal to ZAR 73 091 per DALY averted, a value above the cost-effectiveness threshold of ZAR 38 465.

**Conclusions:** This study indicated that purchasing additional ICU capacity from the private sector may not be a cost-effective use of limited health resources. The ‘real time’, rapid, pragmatic, and transparent nature of this analysis demonstrates a potential approach for further evidence generation for decision making relating to the COVID-19 pandemic response and South Africa’s wider priority setting agenda.

## Background and aim of study

The COVID-19 pandemic has intensified demands on the health care system and resulted in critical shortages of resources (hospital beds, intensive care unit (ICU) beds, ventilators, medical workforce), particularly in the South African public sector. A major area of concern, globally and in South Africa, was the sufficiency of ICU capacity for the management of critically ill COVID-19 patients. Against an ICU bed availability of 3,318 (1,178 public and 2,140 private), the South African Portfolio Committee on Health highlighted a shortfall in ICU beds in the country; where the peak daily demand for ICU beds was projected to be between 4,100 beds (optimistic scenario) and 14,767 (pessimistic scenario).^[1]^ Based on the expected progression of COVID-19 in South Africa and expected utilisation rates of ICU in the management of COVID-19, projections suggested insufficient supply of ICU capacity in the public sector. Government adopted a two-pronged strategy including (1) the adoption of the lockdown strategy to flatten the curve in an effort to reduce the likelihood of demand exceeding the available health care supply and (2) initiatives to purchase critical bed capacity from the South African private sector for use by public sector patients.

Intensive care services are very expensive and are one of the largest drivers of hospital costs, even in public hospitals where the cost per patient per ICU day has been estimated at R22,700.^[2]^ Discovery Health, the largest medical scheme in the country, at the time reported that the average cost of all COVID-19 hospital admissions across its members was R84,708. The average cost of an ICU admission for its members was substantially higher - at R169,525 and these admissions were also reported to have the “highest variation in cost”.^[3]^ Overprovision of ICU beds and subsequent cost escalation through potentially inappropriate use in the private sector was a key finding of the recent Competition Commission Health Market Inquiry.^[4]^

There are a range of health care interventions to manage the progression of the COVID-19 pandemic. Amongst others, resources are required to carry out education, screening, testing, isolation and contact tracing programs, provision of personal protective equipment (PPE) to health workers, treatment in general/high care wards; and in the most critical cases, treatment in ICU. Given the expected downturn in an already weak economy^[5]^ coupled with the increased demand for government resources for economic relief and other measures, the ability of government to commit additional funding to an already under-funded public health sector is limited. Within this context, it is imperative that available public resources are used fairly and efficiently, and that the costs and effects of potential interventions and approaches to care are assessed and weighed against the opportunity costs of their required investment. Traditional economic evaluations can be time consuming with lengthy turnaround times. The rapid pace at which the pandemic unfolded and the imperative it created for policy decisions to be made quickly required the turnaround times for research and analysis informing policy to be shortened substantially. The objective of this study was to assess the cost-effectiveness of ICU management for admitted COVID-19 patients across the public and private health sector in South Africa using a “real time”, pragmatic and transparent health economic modelling approach.

## Methods

### Study design

MOSAIC, a health economic modelling collective established to respond to the need for prompt policy guidance for the South African response to COVID-19, carried out this cost-utility/effectiveness analysis of ICU care. The study was conducted using the principles of the International Decision Support Initiative Reference Case for economic evaluation.^[6]^ It considers two generalised strategies for the inpatient management of severe and critical COVID-19 patients: *(1) general ward and ICU management (GW+ICU)*: admitted patients are managed in a combination of general wards and ICU, with escalation to ICU based on established clinical criteria for severity of disease; (*2) General ward management*: admitted patients are managed in general wards only. Costs are expressed as the cost per case admitted from the health systems perspective, in 2020 South African Rand (ZAR), including both public and private sector costs. Outcomes are expressed as disability adjusted life years (DALYs) and deaths. While the simple measures of “deaths avoided” or “lives saved” are useful and easy to interpret, they miss important treatment effects, such as improvements in morbidity, and they cannot be used to make comprehensive assessments of value for money compared to other treatment options in the health sector. In contrast, representing the impact of interventions as disability-adjusted life years (DALYs) averted allows consideration of health gains due to a reduction of both fatal and non-fatal disease burden; one DALY can be thought of as one lost year of healthy life. Incremental cost-effectiveness ratios (ICERs) are calculated as the difference in costs divided by the difference in health benefits of the treatment strategies, and are compared to a cost effectiveness threshold (CET) derived from an estimate of the marginal productivity of the public health system in South Africa.^[7]^ If the ICER is lower than the defined CET then the marginal opportunity cost of the treatment strategy (in terms of lost health) is expected to be lower than the health benefits of the treatment strategy, indicating that the treatment strategy is likely to represent a cost-effective use of limited resources.^[6]^ The time horizon for the analysis was from admission to discharge or death; while estimates of ongoing morbidity post discharge were included within DALYs, no costs after discharge were estimated. The years of life lost (YLL) from Covid-19 mortality was informed by a secondary actuarial analysis and was not discounted.

### Decision analytic model

A Markov modelling framework was implemented in TreeAge Pro 2020 (TreeAge Software, Inc, Williams-town, Massachusetts, USA) and exported to Microsoft Excel for ease of stakeholder engagement and review, as depicted in Figure 1.

**Figure 1:**
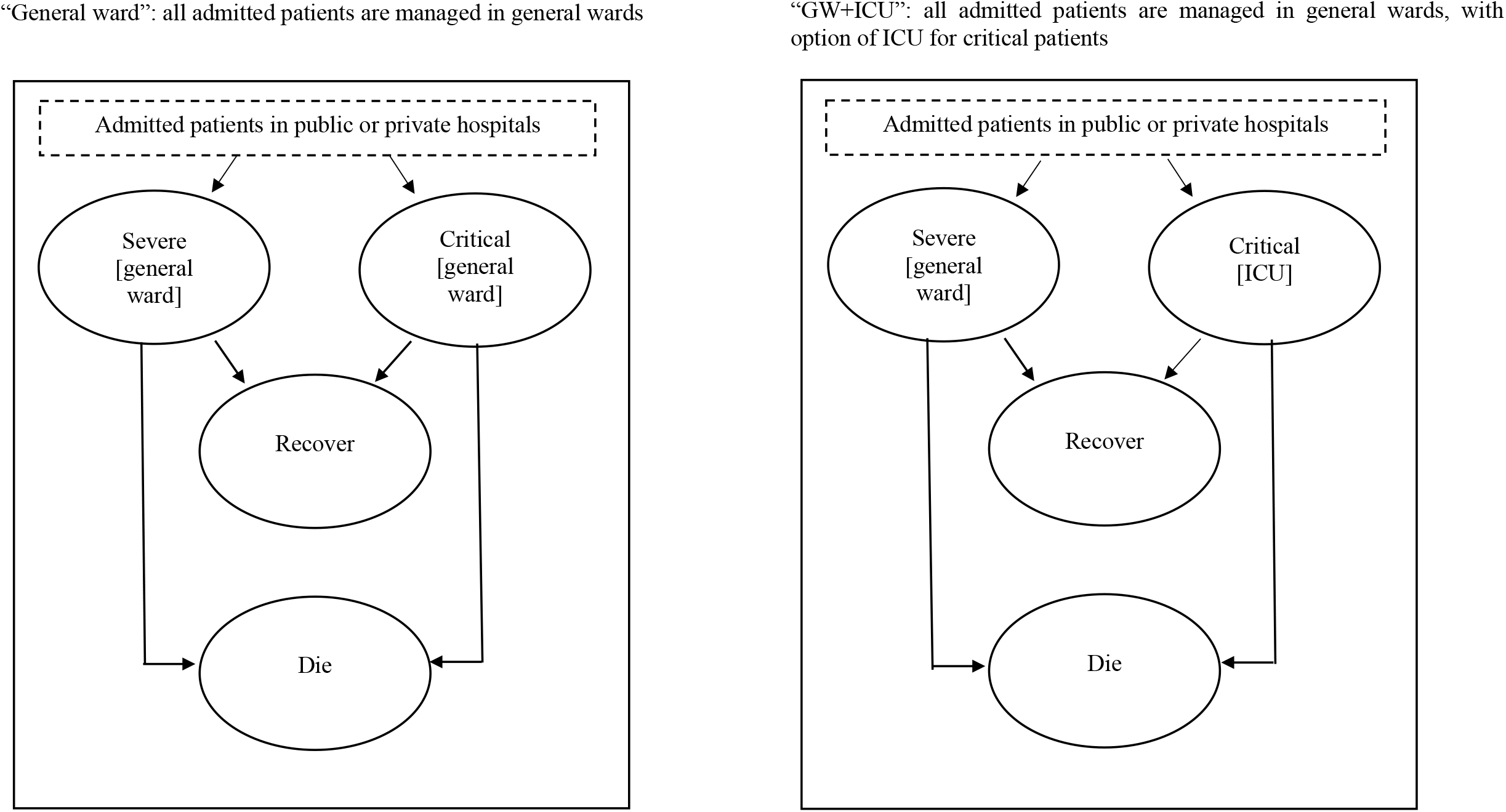
Decision analytic model structure.

The model, available at https://doi.org/10.25375/uct.12382706, runs for a single cycle; which is appropriate to the available secondary data (where estimates are frequently available as rates) as well as the relatively short duration of COVID-19 inpatient care. Patients are ‘randomized’ within the model to each treatment strategy (GW+ICU versus GW); and on admission (to public or private hospitals), patients are modelled as severe or critical. Depending on these factors, patients incur admission costs and accumulate health outcomes as they transition to one of two absorbing states: recover or die. Recovered patients receive a morbidity loss over the duration of their disease and thereafter are assumed to return to their pre-COVID-19 health state while morbidity as well as YLLs are captured for those dying. Further details of these costs and outcomes are provided within Table 1.

**Table 1:** Costs and outcomes in each Markov state.

| Markov state | Costs per Markov state | Transition outcome to recovered | Transition outcome to dead |
| --- | --- | --- | --- |
| Severe patients | Public / private sector cost per hospitalisation in general ward for severe patients | Disability weight for severe patients applied over duration of 1.5 months | Disability weight for severe patients applied over duration of 0.5 months; Years of Life Lost |
| Critical patients | Public / private sector cost per hospitalisation in general ward and ICU for critical patients (GW+ICU model) or general ward only (GW model) | Disability weight for critical patients applied over duration of 2 months | Disability weight for severe patients applied over duration of 0.5 months; Years of Life Lost |

### Model variables

The models rest on 4 different types of variables: *mortality rates* based on severity of illness (i.e. severe versus critical) and approach towards disease management (i.e. GW+ICU versus GW); *utilisation* data including proportions of hospitalized individuals that are critical versus severe, proportion managed in public versus private hospitals and length of stay data for each patient type and management approach; *unit costs* per inpatient day in general wards and intensive care units specific to public and private hospitals; and *DALY* data including YLL, years lived with disability (YLD) and disability weights (DWs). A brief overview of each type of data is provided below. Evidence relating to disease progression and effectiveness of interventions has rapidly changed over the course of the COVID-19 pandemic. The parameters used in the model represent best available evidence as at June 2020.

#### (1) Mortality rates

Mortality rates were extracted from the literature. A systematic search for articles published in English between 01/01/2019 and 30/05/2020 in Medline/PubMed was completed using the terms: “COVID-19” OR “novel coronavirus” OR “SARS-COV-2” OR COVID-19 OR 2019-COV OR “2019 novel coronavirus” AND “clinical characteristics” OR “clinical features” OR “clinical outcomes” AND “death” OR “mortality”. Additional relevant articles were sourced through a manual search of bibliographies of included articles.

As outlined in Figure 2, the results of the initial search were screened by title and abstract. The full texts of potentially relevant articles were retrieved and assessed for inclusion. When articles reported information from the same study sites but at two different time periods, only the articles with the updated statistics were included in this analysis. A total of sixteen observational studies (cross-sectional or cohort) and case series that reported the outcomes of hospitalized COVID-19 patients were included within quantitative synthesis. Average weighted estimates of the case fatality rate among ICU patients and non-ICU patients/patients dying in general ward were calculated using the formula: Deaths/(Deaths + recovered).

**Figure 2:**
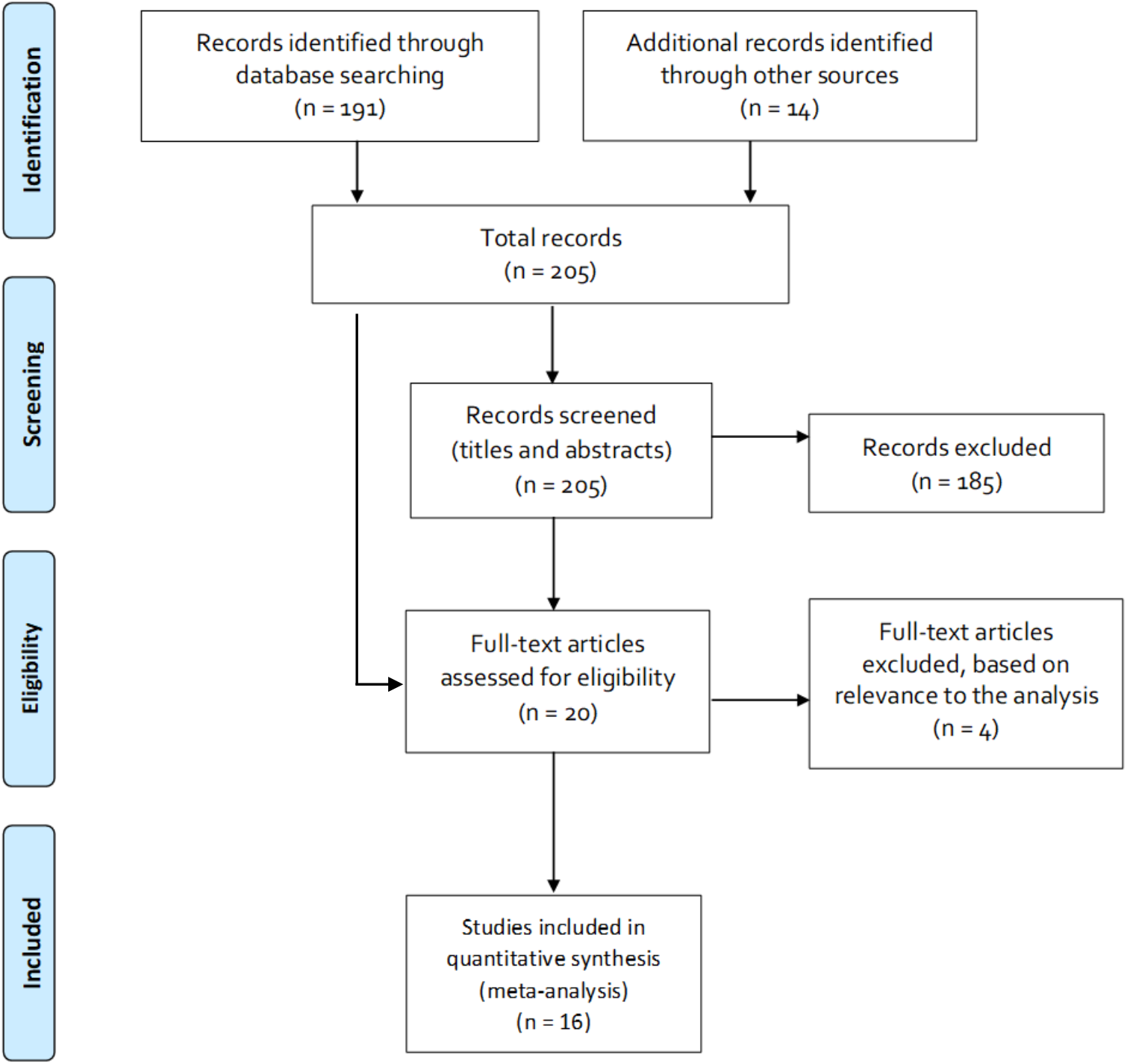
Flow diagram of search strategy and study selection.

#### (2) Utilisation

Utilisation includes the proportions of hospitalized individuals that are critical versus severe and length of stay data for each patient type by type of management (utilization of ICU days by critical patients, utilization of general ward days for severe patients, and for critical patients before/after ICU). These variables were extracted from seven articles^[8-16]^ identified in the above-mentioned systematic search. Average weighted estimates for each variable were calculated. Finally, the proportion of patients using public versus private hospitals was based on the proportion of South Africans with medical scheme membership.^[17]^

#### (3) Unit costs

The model considers the costs of inpatient care in public and private hospitals through the inclusion of unit costs per general ward day and per ICU day. These are multiplied against the abovementioned length of stay estimates to generate a cost per admission. Private sector unit costs are based on the tariff rates in the “Guidelines on Public Private Collaboration in Response to COVID-19” published by the Department of Health.^[18]^ Public sector unit costs were calculated using the Health Systems Trust District Health Barometer (HST-DHB) (12^th^ Edition – 2016/17) datafile^[19]^ which provides hospital-level estimates of expenditure per patient day equivalent (PDE) for all categories of public sector hospitals. These costs were inflated to 2020 prices using the Consumer Price Index^[20]^ and a weighted average unit cost was calculated through weighting unit costs by the percentage of useable beds across levels of care. Because the HST-DHB data do not provide an estimate of the unit cost per ICU day, we estimated this by inflating the average weighted cost by the cost differential between ICU and general ward tariffs in the private sector.

#### (4) DALYs

DALYs are calculated through the summation of YLL and YLD. YLL were informed by a South African actuarial analysis that utilised age- and co-morbidity adjusted mortality rates observed internationally and applied these to the South African population.^[21]^ This resulted in an average estimate of 5.4 YLL per death due to the relatively younger population in South Africa. A wide range of this parameter was tested in sensitivity analysis to reflect the relative uncertainty associated with transferring international mortality data to the South African context. Duration of morbidity is currently unknown for COVID-19; assumptions were therefore made for these parameters. Disability weights for severe/critical COVID-19 patients were based on relevant estimates for similar conditions from the 2017 Global Burden of Disease study.^[22]^

### Sensitivity analysis

Simple sensitivity analyses were run across all variables to assess the impact of changes on the ICER. Where possible, ranges for sensitivity analysis were based on upper and lower confidence intervals, high or low values or interquartile ranges found within the systematic literature review. For the remaining variables, a 50% increase/decrease was implemented, except for where this would move the variable out of feasible range (e.g. mortality rates can only fall within the range 0-1). Thereafter, threshold analyses were run to estimate the percentage change in variables that would render ICU cost-effective, using the published South African cost-effectiveness threshold (CET)^[7]^ as the cut-off for this determination. Finally, an additional scenario was modelled in order to incorporate the effect of administration of the steroid dexamethasone. This analysis entailed the inclusion of the cost of a course of dexamethasone (ZAR 160.85 for twenty 4 mg vials as per 2020 Essential Medicines List price), as well as rate ratio reductions in deaths from ICU (0.65) or from general wards (0.80) as provided in estimates from a UK based randomized controlled trial.^[23]^

### Ethical considerations

This is a modelled cost-effectiveness/utility analysis using published secondary data; no ethical approval was therefore required.

## Results

### Model variables

Table 2 provides a summary of the variables used in the model, together with the ranges on variables used for sensitivity analysis. As highlighted in the table, there are two key unknowns for this economic evaluation that both apply to the GW strategy. The first is the proportion of critical patients dying without access to ICU. This estimate has been based on the high value found in the meta-analysis for critical patients dying from ICU (88% mortality) with ranges for sensitivity analysis including 70% and 100% mortality. The other unknown is the length of stay for these critical patients, which is assumed to be the same as for critical patients managed in ICU.

**Table 2:** Summary of model variables.

| Description | Base Value | Low Value | High Value | Method or assumption (range for sensitivity analysis) | Reference or source |
| --- | --- | --- | --- | --- | --- |
| **Unit costs** |  |  |  |  |  |
| Cost per general ward day public | 3,727.35 | 1,863.68 | 5,591.03 | Average weighted expenditure per patient day equivalent ($\pm 50\%$) | [19] |
| Cost per ICU day public | 17,844.88 | 8,922.44 | 26,767.31 | Average weighted expenditure per patient day equivalent inflated using private tariff differential ($\pm 50\%$) | Assumption |
| Tariff per general ward day private | 5,251.66 | 2,625.83 | 7,877.48 | Published tariff rates ($\pm 50\%$) | [18] |
| Tariff per ICU day private | 25,142.55 | 12,571.28 | 37,713.83 | Published tariff rates ($\pm 50\%$) | [18] |
| **Utilisation** |  |  |  |  |  |
| LoS in general ward in severe patients | 21.25 | 7.25 | 43.00 | Literature review (IQR) | [11, 12] |
| LoS in ICU in critical patients | 8.80 | 4.30 | 13.30 | Literature review (IQR) | [8-10, 13, 14] |
| LoS in general ward in critical patients in absence of ICU | 8.80 | 4.30 | 13.30 | Assumed to be the same as critical patients treated in ICU | Assumption |
| LoS in general ward in critical patients before/after ICU | 1.00 | 0.00 | 3.00 | Literature review (IQR) | [12] |
| Proportion needing ICU | 0.21 | 0.05 | 0.50 | Literature review (low and high value) | [12-15, 24-27] |
| Proportion reliant on public health system | 0.83 | 0.42 | 1.00 | Percentage of population without Medical Scheme coverage ($-50\%$;1) | [28] |
| **Mortality rates** |  |  |  |  |  |
| Proportion of severe patients dying | 0.11 | 0.00 | 0.13 | Literature review (low and high value) | [12, 14-16, 24] |
| Proportion of critical patients dying from ICU | 0.54 | 0.24 | 0.88 | Literature review (low and high value) | [8-10, 12-16, 24, 29, 30] |
| Proportion of critical patients dying without access to ICU | 0.88 | 0.70 | 1.00 | Assumed high value on critical patients dying from ICU (-20%;1) | Assumption |
| **DALYs** |  |  |  |  |  |
| Disability weight in severe patients | 0.13 | 0.09 | 0.19 | Disability weight for severe lower respiratory tract infection (95% CI) | [22] |
| Disability weight in critical patients | 0.41 | 0.27 | 0.56 | Disability weight for severe pneumoconiosis (95% CI) | [22] |
| Duration of illness in severe patients | 0.13 | 0.06 | 0.19 | 1.5 months (±50%) | Assumption |
| Duration of illness in critical patients | 0.17 | 0.08 | 0.25 | 2 months (±50%) | Assumption |
| Duration of illness prior to death | 0.04 | 0.02 | 0.06 | 0.5 months (±50%) | Assumption |
| Years of life lost if dying from Covid-19 | 5.40 | 2.70 | 8.10 | 5.4 years (±50%) | [21] |
| **Other** |  |  |  |  |  |
| Cost-effectiveness threshold per DALY averted | 38,465.46 |  |  | Used to assess value for money; if ICER<CET intervention potentially cost-effective | [7] |

Other important unknowns include disability weights for COVID-19, with the DW for severe lower respiratory tract infection assumed for severe COVID-19 patients and the DW for severe pneumoconiosis assumed for critical patients, as extracted from the global burden of disease study.^[22]^ Duration of illness is assumed to be 0.5 months in those dying, 1.5 months in severe patients and 2 months in critical patients. Finally, YLL is based on international mortality rates per 100 000 population by age and comorbidity applied to the South African demographic structure.^[21]^

### Cost-effectiveness

Table 3 summarizes the cost-effectiveness results. Assuming base case values across all variables, the model produces a cost per admission of ZAR 75 127 versus ZAR 103 030, deaths at 27% versus 20%, and DALYs at 1.48 versus 1.10 in the general ward versus ICU strategy, respectively. The incremental cost-effectiveness ratio is ZAR 73 091 per DALY averted or ZAR 390 798 per death averted. The general ward strategy requires 18.85 general ward days per admission, while the ICU strategy requires 17 general ward days and 1.85 ICU days.

**Table 3:** Base case cost-effectiveness results (2020 ZAR)

| Strategy | Cost (ZAR) | DALYs | Deaths | ICU Days | Genl. Ward Days | Incr. Cost per DALY Averted | Incr. Cost per Death Averted |
| --- | --- | --- | --- | --- | --- | --- | --- |
| Genl. Ward | 75 127.25 | 1.48 | 0.27 | - | 18.85 |  |  |
| Genl. Ward + ICU | 103 030.20 | 1.10 | 0.20 | 1.85 | 17.00 | 73 091.37 | 390 797.60 |

DALYs: disability adjusted life years; Genl.: general; ICU: intensive care unit; Incr.: incremental.

### Sensitivity analysis

Figure 3 presents the tornado diagram generated from the simple sensitivity analyses, depicting the seven analyses that generated the largest changes to the ICER. These results are compared against the cost-effectiveness threshold of ZAR 38,465 per DALY averted [7] to assess whether changes in variables would render ICU cost-effective.

**Figure 3:**
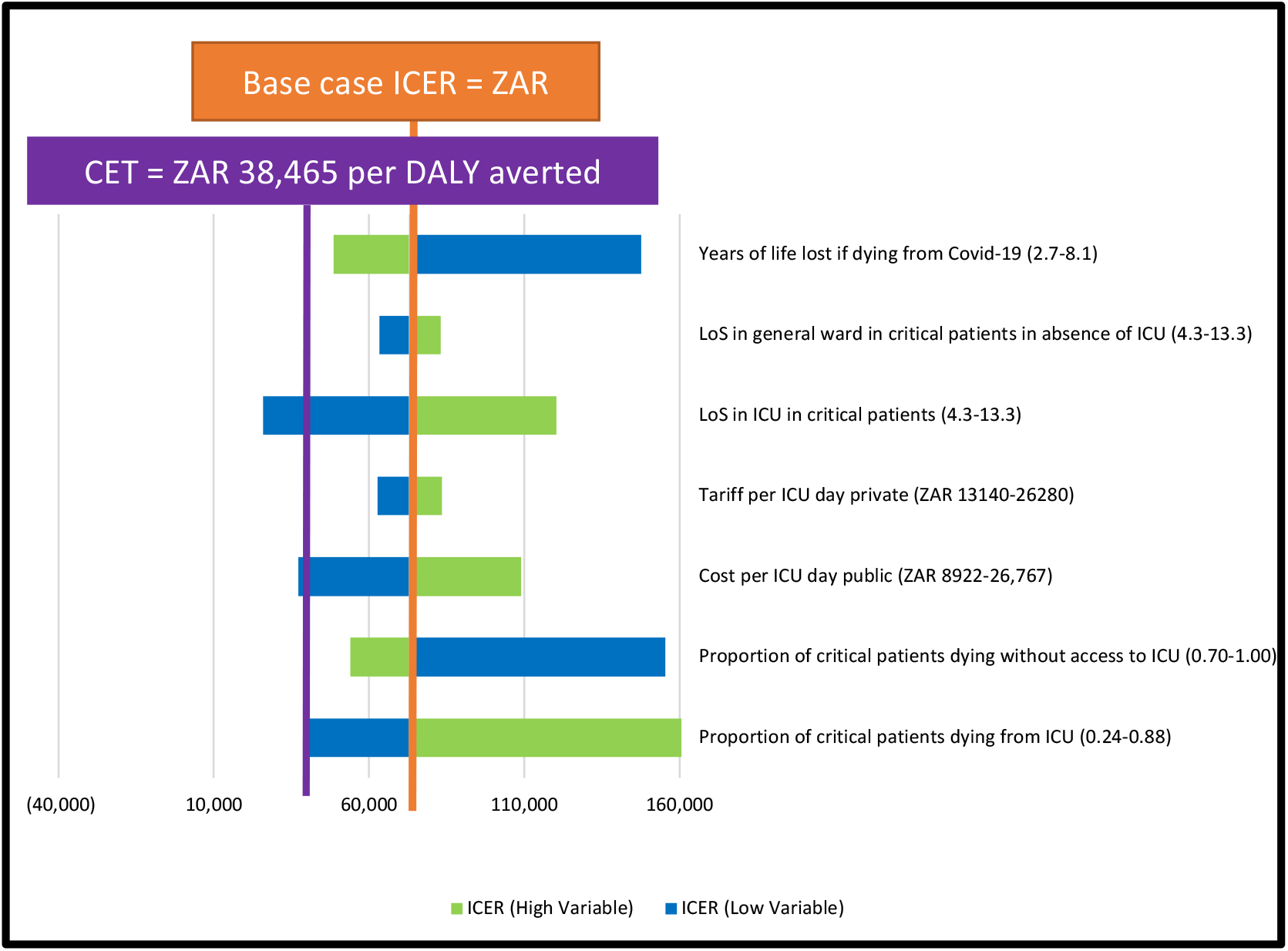
Tornado diagram summarizing simple sensitivity analyses.

As is shown in Figure 3, three analyses showed promise for generating cost-effective ICERs: (1) length of stay in ICU in critical patients; (2) cost per ICU day in the public sector; and (3) proportion of critical patients dying from ICU. In effect, these analyses indicate that if ICU were less costly (either through reduction in the unit cost or the length of stay) or more effective, intensive care would be more likely to be a cost-effective use of scarce health system resources.

The threshold analysis (Table 4) takes this one step further to estimate the exact percent change in variables needed to generate cost-effective ICERs. As is shown, five threshold values were found. These thresholds included a 57% decrease in the proportion of critical patients dying from ICU; 48% decrease in the public sector cost per ICU day; 38% decrease in ICU length of stay; 179% increase in LoS in critical patients managed in general wards; and 89% increase in YLL in those dying from COVID-19. Finally, the scenario analysis on the inclusion of dexamethasone generates a slightly improved ICER of ZAR 70,400 per DALY averted.

**Table 4:** Threshold analyses.

| **Variable Description** | **Base Value** | **Threshold Value** | **% Change** | **Interpretation** |
| --- | --- | --- | --- | --- |
| Proportion of critical patients dying from ICU (pdeadICU) | 0.54 | 0.23 | -57% | 57% decrease in ICU mortality renders ICU cost-effective |
| Cost per ICU day public (cICUpub) | 17,845 | 9,227 | -48% | 48% decrease in cost per ICU day in public sector renders ICU cost-effective |
| LoS in ICU in critical patients (uICU) | 8.80 | 5.5 | -38% | 38% decrease in ICU length of stay renders ICU cost-effective |
| LoS in general ward in critical patients in absence of ICU (uIPDcritical) | 8.80 | 24.59 | 179% | 179% increase in length of stay for critical patients managed in general ward renders ICU cost-effective |
| Years of life lost if dying from COVID-19 (YLL) | 5.40 | 10.21 | 89% | 89% increase in YLL in those dying renders ICU cost-effective |

## Discussion

Based on the evidence available at the time and at base case values, this economic evaluation indicated that expanding ICU capacity through purchasing from the private sector may not be a cost-effective use of scarce health system resources. While intensive care is expensive, if it were effective at preventing deaths in critically ill COVID-19 patients, it would have generated more favourable results. However, our meta-analysis of evidence from nine available studies suggested that outcomes were poor ^[8-16, 24, 29, 30^]. The mortality rate for critically ill patients managed in ICU was on average 54% (range: 28%-88%) meaning that for a substantial proportion of critical COVID-19 patients, admission to ICU was unlikely to be a life-saving intervention. However, if the cost per admission could be reduced (e.g. through reducing lengths of stay in ICU), if outcomes were to improve, or if ICU were targeted at patients with higher potential years of life to lose, it would become more cost-effective. While the inclusion of dexamethasone improved the ICER results, it did not move the ICER into the cost-effective range. That said, the dexamethasone scenario illustrates how the inclusion of new technologies such as medicines could generate changes in the economics of inpatient care for COVID-19.

Approaches to decision making related to allocation of resources in the face of scarcity (also called “priority setting”) commonly includes four fundamental values or ethical principles drawn from theories of distributive justice:[^31, 32^] (1) “utilitarianism” - doing the greatest good for the greatest number of people, either by saving the highest number of lives and/or saving the largest number of life-years, which is the basis of cost-effectiveness; (2) “egalitarianism” – providing equal access or equal treatment for equal need; (3) “rule of rescue” – providing urgent, life-saving treatments irrespective of the cost; and (4) “desert” - promoting and rewarding social usefulness.^[33]^ Based on utilitarian or egalitarian approaches, there would be limited merit in government investing heavily in ICU capacity. However, justification for investing in ICU may be found through the application of other ethical principles such as the “rule of rescue”. The latter calls on society to respond to the extreme risk faced by an identifiable individual, however it generally only holds when the numbers are small. The difficulty to face with COVID-19 is that while many of us align with a rule of rescue based response, the stark realities of resource constraints call on us to acknowledge that the allocation of resources to expand ICU capacity (1) would be unsustainable given the costs and (2) would lead to a greater loss of life through the crowding out of more cost-effective health services.

While the economic evaluation methodology can provide quantitative evidence to inform each of these ethical principles, and hence is a key input to any consideration of distributive justice,^[34]^ experience globally suggests that a multi-ethical framework is more likely to result in a fairer allocation of resources. Moreover, because reasonable people should be expected to disagree on the relative merits of these ethical principles, priority setting needs to be vested within procedural justice, a key aspect of which is transparent deliberation.^[35]^ Our real-time and open access approach to modelling provides one example of how such transparency can be facilitated.

There are a number of limitations to this study and similar studies. The urgency to inform decision making and restrictions on primary data collection necessitated a reliance on secondary data while the ongoing emergence of new information required flexibility in model building. To address these concerns (1) a comprehensive systematic review of the available evidence was carried out to ensure that all of the available information was fed into the model and (2) an open access modelling framework^[36]^ with a user guide^[37]^ was developed to facilitate full exploration of uncertainty through sensitivity analysis and to allow for parameters to be quickly and easily updated as new information becomes available.

## Conclusion

This rapid analysis provided key evidence on the cost-effectiveness of alternative management strategies in the hospital care of critical and severe patients with COVID-19 disease in the South African setting. It has shown that ICU use for COVID-19 patients was unlikely to be cost-effective on the margin, and therefore an expansion of ICU capacity through purchase of private sector capacity for use by public sector patients (at current prices and evidence of effectiveness) may not be the best use of limited health resources, whether from utilitarian or egalitarian ethical perspectives.

There are few (if any) examples of decision analytic modelling and cost effectiveness analysis being conducted in “real time” to inform policy decisions in the South African public health sector. The rapid, pragmatic and transparent analysis employed by the MOSAIC group demonstrates a potential approach for further evidence generation for decisions relating to the COVID-19 pandemic response and South Africa’s wider priority setting agenda.

## Data Availability

Data are available online

https://doi.org/10.25375/uct.12382706

